# Implementing community-based minimally invasive tissue sampling for mortality surveillance in rural South Africa: A process evaluation of the Vukamberi project

**DOI:** 10.64898/2026.11.08.26367567

**Authors:** Zokwane Lucky Mondlane, Kathleen Kahn, Shingirai Chipungu, Ryan Gregory Wagner, Beth Tippett Barr, Jessica Price

## Abstract

Accurate cause-of-death data are limited in rural low- and middle-income settings, where most deaths occur at home without medical certification. Minimally invasive tissue sampling (MITS) is a culturally acceptable alternative to complete autopsy but has largely been applied to stillbirths and child deaths. We evaluated the process and implementation of a community-based, all-age MITS study in rural South Africa.

The Vukamberi project (“to examine” in Xitsonga) is a prospective study in rural Bushbuckridge sub-district, Mpumalanga Province. Guided by the Medical Research Council Framework for Complex Interventions and Proctor’s Implementation Outcomes Framework, we assessed reach, adoption, fidelity, feasibility, acceptability and timeliness using prospectively collected records of deaths notified between 07 July 2024 to 30 June 2026, and mapped challenges to adaptive strategies by domain and phase.

All 408 notified community deaths were screened, and 316 (77.5%) were eligible. Families consented for 206 eligible cases (65.2%), all of which proceeded to MITS, with target-organ sampling adherence in 199 (96.6%). Median time from death to notification was 15 hours (interquartile range [IQR] 5–23) and to MITS completion 37 hours (IQR 25–49). Enrolled decedents were 197 adults (95.6%) and 9 paediatric cases (4.4%). Main ineligibility reasons were in-hospital death (excluded by design; 27 cases) and notification beyond 48 hours (26 cases). Family refusal, mostly reflecting grief-related unreadiness and sociocultural barriers, accounted for 53 (48.2%) non-enrolled eligible cases. Adaptive strategies that increased enrolment included extending recruitment beyond the Agincourt Health and Socio- Demographic Surveillance System into the wider sub-district, incentivised community informants, elder-mediated consent, gender-appropriate field staff, non-disfigurement messaging in local languages and mortuary-based consenting where household access was restricted by a traditional leader.

Community-based MITS was feasible in this rural setting. Culturally responsive consent strategies, mortuary partnerships and multi-channel death notification helped overcome sociocultural and logistical barriers, and a reliable supply of consumables proved essential for continuous enrolment. These findings may inform community-based mortality surveillance in other low- and middle-income settings.

## Introduction

Accurate and timely cause-of-death (CoD) data are critical for informing public health decision- making, resource allocation, and health-system accountability [1]. In many low- and middle-income countries (LMICs), a substantial proportion of deaths occur outside health facilities and without medical certification, resulting in inaccurate data for national statistics and global burden of disease estimation, and inadequate data to guide policies aimed at reducing mortality [2,3].

Sub-Saharan Africa carries a disproportionately high burden of unregistered community deaths, compounded by underdeveloped civil registration and vital statistics (CRVS) systems [4,5]. Delays and incompleteness in death reporting, compounded by logistical barriers, contribute to poor-quality mortality data [4,5]. These gaps limit the ability of local governments and global health decision-makers to predict and detect emerging outbreaks, monitor progress towards Sustainable Development Goals (SDGs), and understand the definitive burden of infectious and non-communicable diseases (NCDs) contributing to mortality [6,7].

South Africa presents a paradox in mortality reporting; despite relatively high national CRVS coverage, substantial subnational heterogeneity persists, particularly in rural settings where deaths occur at home, medical certification of cause of death (MCCOD) is less accessible, and sociocultural norms influence death reporting [8]. In rural settings, most deaths that occur at home are witnessed by family members, and verbal autopsy (VA) has thus become a widely used tool to determine the probable cause-of-death [9]. Although VA is a valuable, pragmatic tool for understanding population-level mortality patterns, especially in settings with incomplete civil registration systems, it has several limitations, primarily when applied to individual-level diagnosis or detailed clinical analysis [10].

Minimally invasive tissue sampling (MITS) has emerged in the last decade as a culturally acceptable alternative to complete diagnostic autopsy (CDA) [11,12]. MITS uses needle-based biopsy sampling of key target organs, and collection of cerebrospinal fluid, blood and nasopharyngeal swabs, enabling microbiological, molecular, and histopathological evaluation with substantially improved ability to assess cause of death [13,14]. The Child Health and Mortality Prevention Surveillance (CHAMPS) network has demonstrated that MITS can identify infectious causes of death, clarify causal pathways, and quantify the burden of preventable mortality in LMICs [15,16]. Studies have also shown that MITS achieves high community acceptability when implemented with culturally aligned engagement strategies, transparent consent processes, and return of results to families of the deceased [17,18].

However, the global MITS evidence base is still dominated by facility-based MITS and child mortality surveillance. Far less is known about conducting community-based MITS in rural settings across all age-groups, where logistical, operational, and social constraints differ markedly [18]. In addition to the time constraints in which to conduct MITS after death, community-based deaths pose unique challenges, including rapid burial expectations driven by cultural and religious practices, geographically dispersed families, delayed death notification to the study team, multigenerational decision-making involving elders, and mistrust or uncertainty about post-mortem procedures [19].

Community-based MITS implementation evidence on operational workflow, barriers, facilitators, and adaptive strategies is limited [20]. Implementation science frameworks show that evidence essential for translating effective interventions into practice encompasses contextual factors, particularly the acceptability, feasibility, and fidelity of the intervention in real-world settings [21]. Community-based surveillance in the region rests on volunteer notification networks whose structures and knowledge levels vary widely [22], and there is correspondingly limited knowledge on how community-based MITS can be systematically integrated into rural mortality surveillance platforms, how notification systems can be strengthened to minimise delays, and how cultural and social institutions such as traditional leadership, elders, burial societies and mortuary partnerships can be leveraged to influence implementation success.

We address this gap through a structured process evaluation of the Vukamberi MITS project (“to examine” in Xitsonga). We report key process and implementation outcomes (reach, adoption, fidelity, feasibility, acceptability and timeliness), the operational and sociocultural barriers encountered, and the adaptive strategies used to address them, and early operational and laboratory findings, to inform scalable approaches to community mortality surveillance.

## Materials and methods

### Study design

We conducted a systematic process evaluation of the Vukamberi MITS Project, measuring implementation outcomes including notification timelines, consent rates, procedural completion, and sample adequacy, on all deaths recruited from 07 July 2024 to 30 June 2026. The Vukamberi MITS protocol is the intervention being evaluated and is summarised narratively below, while the Agincourt HDSS is the platform within which the intervention was delivered and is summarised in the following section, with a full description in S1 Appendix. The study protocol has not been published and is available from the corresponding author on request.

### Study setting

#### Agincourt HDSS

The Vukamberi MITS project is implemented in the Agincourt Health and Demographic Surveillance System (HDSS), though recruitment extended beyond the boundaries of the HDSS into the wider Bushbuckridge sub-district in Mpumalanga province, north-east South Africa. The HDSS is a South African Medical Research Council/Wits research platform that has followed a rural population of approximately 120 000 people for over three decades, recording every birth, death and migration and completing a verbal autopsy on each recorded death [23]. Three features of this setting bore directly on implementation: most deaths occur at home and outside clinic hours, when medical attendance is rare; high rates of circular and temporary labour migration frequently placed the consenting next-of-kin away from the household within the 48-hour operational window; and from August 2025 a local traditional leader restricted study access to 14 villages in the catchment, which determined where families in those villages could be approached for consent. The HDSS population, the wider sub-district and its health facility network are described in full in S1 Appendix the study area is shown in Fig 1.

**Fig 1.**
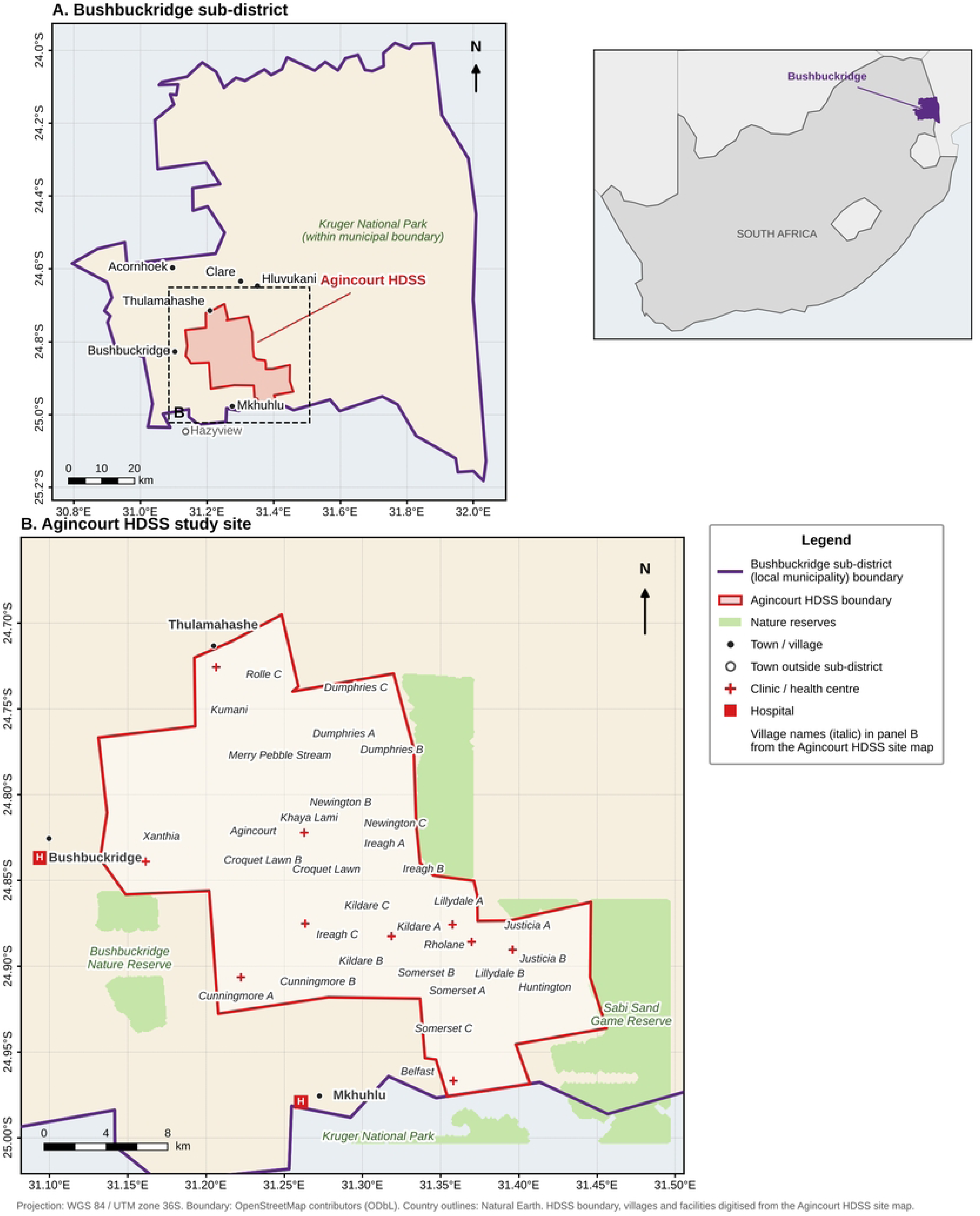
Study area. (A) Bushbuckridge sub-district (local municipality), Mpumalanga Province, South Africa, showing the location of the Agincourt HDSS; inset shows the sub-district within South Africa. (B) Agincourt HDSS villages, health facilities and adjacent nature reserves. Map produced in Python (GeoPandas) in WGS 84 / UTM zone 36S. Municipal boundary from OpenStreetMap (© OpenStreetMap contributors, ODbL); country outlines from Natural Earth; the HDSS boundary and village and facility locations were digitised and georeferenced from the Agincourt HDSS site map.

#### Vukamberi MITS

The Vukamberi project is an ongoing, prospective MITS study that began with a qualitative study from May to December 2023 on key stakeholders to better understand community perspectives and potential facilitators and barriers to the introduction of MITS [24], and then transitioned into an ongoing intervention study involving the activation of the Research Triangle Institute (RTI) MITS protocol and enrolment of community deaths across all ages. In operational terms the protocol proceeds as follows: community deaths are communicated to the study team through a multi-channel network of incentivised community key informants, mortuary partners and primary health care clinics [25], with a target of notification within 48 hours of death. Deaths were eligible if they occurred outside a health facility (or the decedent was declared dead on arrival), had been notified within 48 hours of death, were of natural causes, occurred to a resident of Bushbuckridge sub-district, and had a family member able to give written informed consent within the 48-hour window. Field staff screen every notified death against the eligibility criteria. For eligible deaths, one male and one female community- facing field worker jointly approach the family, in Xitsonga, Sepulana (Northern Sotho) or English, to obtain written informed consent, either at the household or, where household access is restricted or preferred by the family, at the mortuary; elder household members are engaged as consent mediators where appropriate. Consented cases are sampled by a trained MITS team using standardised percutaneous needle biopsy of brain, lungs and liver, and of kidneys where field time allows (attempted for all cases rather than selected by cause of death, given its relevance to the post-mortem kidney sub-study), together with cerebrospinal fluid, blood and nasopharyngeal swabs, following MITS Surveillance Alliance standard operating procedures, with point-of-care serological testing performed at the time of sampling. Specimens are transported to the in-house Agincourt laboratory for microbiology, with histopathology referred externally. Results are combined with verbal autopsy and, where available, ante-mortem clinical records at a multi-disciplinary Determination of Cause of Death (DeCoDe) panel. The cause of death is then returned in person to the family. This workflow is shown in Fig 2.

**Fig 2.**
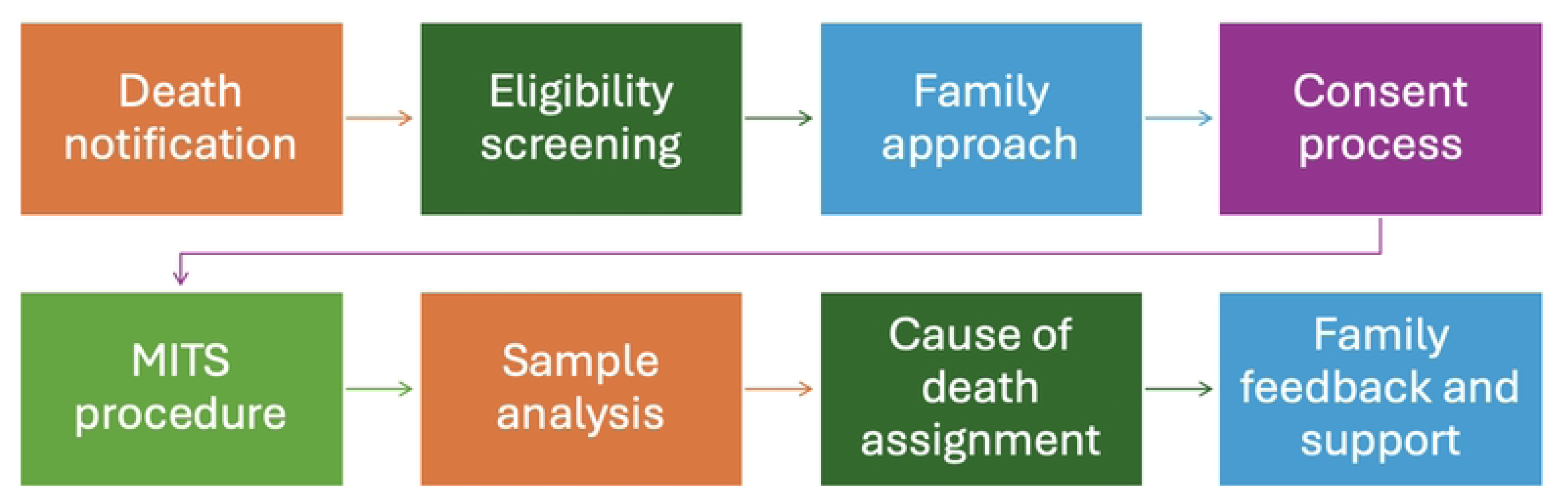
Operational workflow for community-based MITS in the Vukamberi project.

### Process evaluation framework

The Vukamberi project was assessed using a process evaluation framework integrating the Medical Research Council (MRC) Framework for Developing and Evaluating Complex Interventions [26], with Proctor’s Implementation Outcomes Framework [27]. The MRC framework guided assessment of the intervention context, implementation mechanisms, and pathways through which inputs (community engagement, mortuary partnerships, trained field teams, laboratory capacity) enabled delivery of the community-based MITS workflow. Seven implementation outcome domains were specified for this evaluation. Four are Proctor constructs (adoption, fidelity, feasibility and acceptability). Reach, timeliness and adaptations were added from complementary sources to capture features specific to community-based post-mortem surveillance. Reach was taken from the RE-AIM framework [28] and corresponds to Proctor’s penetration construct; timeliness is a core attribute in the CDC guidelines for evaluating public health surveillance systems [29] and was included because MITS must be performed soon after death, before decomposition compromises sample quality; and adaptations were documented as recommended by the MRC framework, which treats adaptation to context as central to complex intervention delivery [26]. The seven domains were operationalised as follows: reach (timely death notification and eligibility screening), adoption (family consent and enrolment), fidelity (adherence to MITS standard operating procedures), feasibility (procedural completion and sample adequacy), acceptability (cultural appropriateness of the process, including elder involvement in consent), timeliness (intervals from death to notification and sampling), and adaptations (modifications introduced in response to operational and sociocultural barriers). This integrated approach enabled assessment of whether community-based MITS was feasible and of how specific mechanisms and contextual factors shaped implementation outcomes across phases (Fig 3).

**Fig 3.**
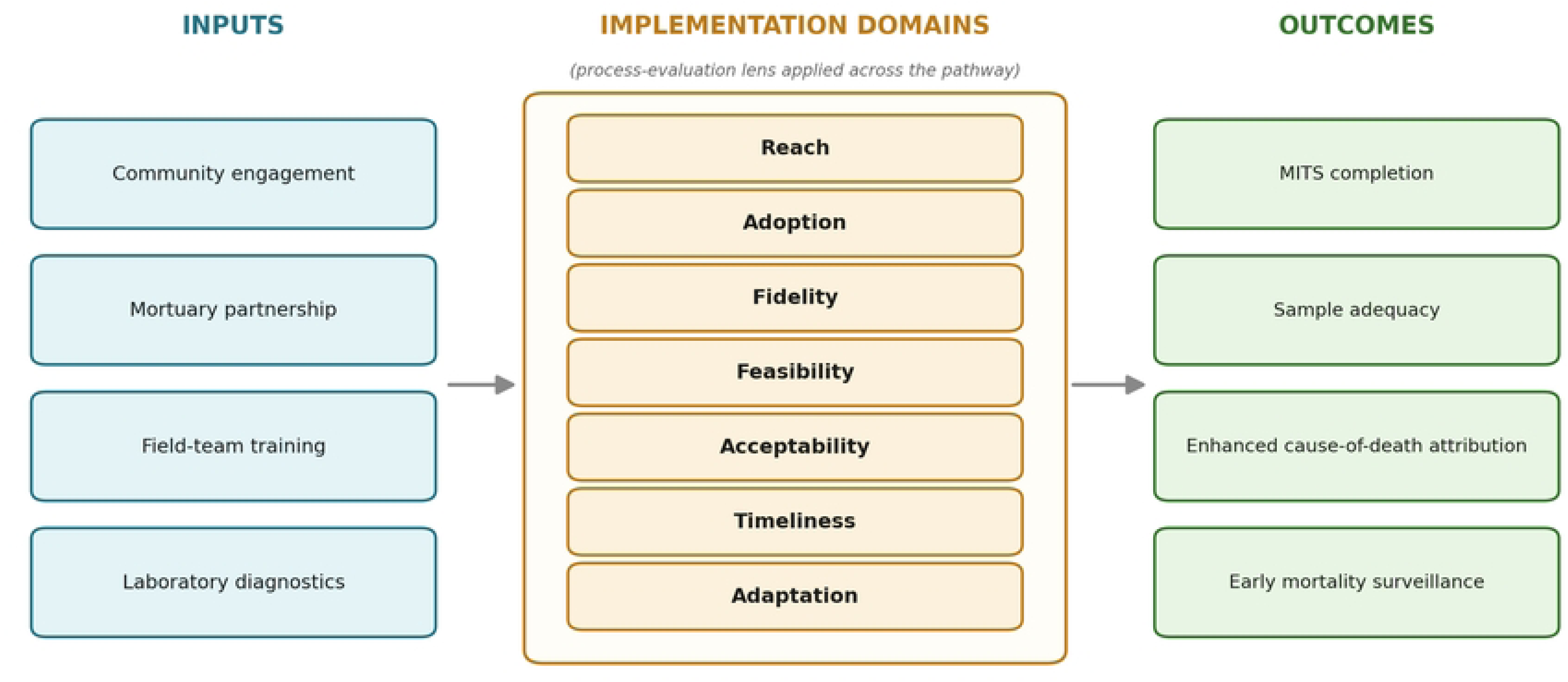
Process evaluation framework for community-based MITS implementation.

### Data collection

Data were collected prospectively from study-specific operational and implementation records, including monthly death notification logs; field team operational and case reports; consent documentation forms; in-house laboratory sample quality records and results; and monthly surveillance summary reports compiled by the study team. Implementation challenges and adaptive responses were documented during weekly team review meetings and mapped to implementation outcome domains.

### Data analysis

The operational phases were sequential, and our assessment of each of Proctor’s implementation domains extends across multiple phases. The remainder of the results are therefore discussed thematically under each outcome domain namely: (1) death notification and screening (reach and timeliness); (2) eligibility assessment; (3) consent and enrolment (adoption and acceptability); (4) procedural completion and sample collection (fidelity and feasibility); and (5) laboratory analysis status.

Quantitative implementation domains (reach, adoption, fidelity, feasibility, acceptability, and timeliness), drawn from Proctor’s Implementation Outcomes Framework [27] and the complementary sources described above, were summarised using descriptive statistics, including frequencies, proportions, medians with interquartile ranges (IQR), and ranges where appropriate. Enrolment trajectories were analysed monthly to characterise programme ramp-up and phase-specific performance. Implementation challenges and adaptive strategies were categorised as operational, logistical, sociocultural, or project-level, and mapped against the implementation domains and the phase in which they occurred, enabling a temporal analysis of barrier-adaptation relationships. To describe change in implementation performance over time, the reach, adoption and feasibility indicators were additionally disaggregated by implementation year (Year 1: July 2024–June 2025; Year 2: July 2025–June 2026), and the enrolment cascade was tabulated cumulatively by quarter. The protocol was open to decedents of all ages from inception, and the age distribution of the enrolled series was described to establish whether reach extended across age groups. Age groups followed the study protocol: stillbirth, neonate (under 28 days), infant (28 days to under 1 year), child (1– 16 years) and adult (older than 16 years).

### Ethical approval

Ethical approval was obtained from the University of the Witwatersrand Human Research Ethics Committee (M230754 MED23-07-051). The study was conducted in accordance with the Declaration of Helsinki and the Protection of Personal Information Act 4 of 2013 (POPIA). Written informed consent was obtained from the next of kin of each enrolled decedent prior to any study procedures whom aged 18 years and above. Participation was voluntary, and emotional and psychological support was offered to all bereaved families during and after the consent process.

## Results

Implementation proceeded through four phases: a preparatory phase before enrolment began, followed by a pilot, scale-up, and full surveillance across the 24-month period. The preparatory phase (May–December 2023) established community and stakeholder engagement, formative acceptability work and ethics approval. The pilot phase (July–December 2024) tested the standard operating procedures, established the community informant network and introduced mortuary-based consenting. Scale-up (January–December 2025) extended the catchment area beyond the HDSS, incentivised community informants and formalised mortuary partnership agreements. And finally, the full surveillance phase (January–June 2026) established the in-house culture bench, initiated the verbal autopsy and DeCoDe workflows, expanded the field team from one to two community liaison officers with an additional MITS assistant, and introduced buffer stock protocols following the kit supply gap. Screening coverage was completed every month, while enrolment rose from fewer than five procedures per month in the pilot to 12–26 per month from June to November 2025 and, after a commodity stockout in early 2026, 14–15 per month from April to June 2026. The major adaptations introduced at each phase, and the barriers they were designed to overcome, are summarised in Table 1.

**Table 1.** Implementation phases and key adaptations to overcome operational, sociocultural, and logistical barriers to community-based MITS, Vukamberi project, May 2023 – June 2026.

| Phase | Period | Key activities | Major adaptations introduced |
|---|---|---|---|
| 1: Preparatory | May–Dec 2023 | Community engagement, stakeholder mapping, acceptability work, ethics approval | Public Engagement Office (PEO) involvement; Community Advisory Board consultations; formative sociocultural assessment |
| 2: Pilot rollout | Jul–Dec 2024 | Pilot enrolment of community deaths; Standard Operating Procedure (SOP) testing; notification system establishment | Mortuary consenting introduced; community informant network established; elder engagement protocols developed |
| 3: Scale-up | Jan–Dec 2025 | Expansion of notification network and catchment area; | Informant incentivisation; catchment area expansion; formal mortuary partnership agreements |
| 4: Full surveillance | Jan–Jun 2026 | Laboratory scale-up; DeCoDe panel initiation; field team expanded from one to two community liaison officers, with an additional MITS assistant recruited, increasing capacity for notification follow-up, consent engagement, and sample collection | In-house culture bench established; VA and DeCoDe workflows initiated; buffer stock protocols introduced after kit supply gap |

### Death notification and screening

Between July 2024 and June 2026, 408 community deaths were communicated to the Vukamberi project surveillance system within the study catchment area. All 408 deaths (100%) were screened for MITS eligibility, demonstrating complete screening coverage across all 24 months of the enrolment period. This was achieved through a multi-channel notification infrastructure integrating incentivised community informants and mortuary partnerships. Screening coverage was maintained at 100% throughout, although only 6 deaths were reported in February 2026, when just two mortuaries reported to the study team; screening of all notified deaths was maintained, but ascertainment of deaths occurring in that month was incomplete. Among the 180 enrolled cases with valid dates and times of death and notification, median time from death to notification was 15.2 hours (IQR 5.2–23.0). Implementation performance across all outcome domains is summarised in Table 2.

**Table 2.** Implementation performance indicators for community-based MITS, Bushbuckridge sub-district, July 2024 – June 2026.

| **Outcome** | **Indicator definition** | **Overall N, %** | **Year 1 Jul 2024 – Jun 2025** | **Year 2 Jul 2025 – Jun 2026** |
|---|---|---|---|---|
| **Reach** | Proportion of notified deaths screened for eligibility | 408/408, 100% | 182/182, 100% | 226/226, 100% |
| **Reach** | Proportion of screened deaths meeting eligibility criteria | 316/408, 77.5% | 119/182, 65.4% | 197/226, 87.2% |
| **Adoption** | Consent rate among eligible deaths | 206/316, 65.2% | 70/119, 58.8% | 136/197, 69.0% |
| **Fidelity** | SOP* adherence for target organ sampling | 199/206, 96.6% | N/A | N/A |
| **Feasibility** | Procedural completion among consented cases (% completion) | 206/206, 100% | 70/70, 100% | 136/136, 100% |
| **Sample adequacy** | Adequate samples across all target organs | Brain – 203/206 (98.5%)<br>Liver – 204/206 (99.0%)<br>Right lung – 205/206 (99.5%)<br>Left lung – 204/206 (99.0%)<br>Additional: Right kidney – 24/206 (11.7%) and left kidney – 29/206 (14.1%) | N/A | N/A |
| **Acceptability** | Proportion of consent encounters involving elder household members | 277/316 (87.7%) | N/A | N/A |
| **Timeliness** | Time from death to MITS completion (median, IQR) | 37.3 h (24.5–48.9); n = 198 | N/A | N/A |

\*SOP = standard operating procedure. Year 1 is July 2024 – June 2025 and Year 2 is July 2025 – June 2026; counts are of deaths notified within each period. N/A = not applicable and marks an indicator recorded at case level and not captured in the monthly surveillance summaries, which is therefore reported for the full period only.

### Eligibility assessment

Of 408 deaths screened, 316 (77.5%) met the eligibility criteria for MITS. The remaining 92 deaths (22.5%) were assessed as ineligible. Cumulative eligibility was 81.0% in the first quarter (July–September 2024), fell to 48.3% by the end of the pilot (December 2024) and then rose in every subsequent quarter to 77.5% by June 2026 (Table 3), reflecting operational strengthening of the notification and response system.

**Table 3.** MITS enrolment cascade (cumulative) by quarter, Bushbuckridge sub-district, July 2024 – June 2026.

| **INDICATORS** | **Q3: 2024**<br>**Jul–Sep** | **Q4: 2024**<br>**Oct–Dec** | **Q1: 2025**<br>**Jan–Mar** | **Q2: 2025**<br>**Apr–Jun** | **Q3: 2025**<br>**Jul–Sep** | **Q4: 2025**<br>**Oct–Dec** | **Q1: 2026**<br>**Jan–Mar** | **Q2: 2026**<br>**Apr–Jun** |
|---|---|---|---|---|---|---|---|---|
| **Deaths notified (cumulative)** | 21 | 58 | 106 | 182 | 256 | 305 | 342 | 408 |
| **Screened (n, %)** | 21 | 58 | 106 | 182 | 256 | 305 | 342 | 408 |
| **New deaths** | 21 | 37 | 48 | 76 | 74 | 49 | 37 | 66 |
| **Eligible (n, % of cumulative deaths)** | 17<br>(81.0%) | 28<br>(48.3%) | 65<br>(61.3%) | 119<br>(65.4%) | 186<br>(72.7%) | 230<br>(75.4%) | 264<br>(77.2%) | 316<br>(77.5%) |
| **Eligible, not enrolled (n, cumulative)** | 4 | 10 | 28 | 49 | 64 | 76 | 102 | 110 |
| **of which family refusal (n, cumulative)** | 1 | 3 | 18 | 29 | 36 | 45 | 46 | 53 |
| **MITS done (n, % of cumulative eligible deaths)** | 13<br>(76.5%) | 18<br>(64.3%) | 37<br>(56.9%) | 70<br>(58.8%) | 122<br>(65.6%) | 154<br>(67.0%) | 162<br>(61.4%) | 206<br>(65.2%) |

Cumulative totals are reported at the end of each period. Eligibility % is calculated against cumulative deaths notified; MITS done % is calculated against cumulative eligible deaths. Eligible deaths are the sum of MITS done and eligible deaths not enrolled. The three non-enrolments without a recorded reason are assigned to August 2024, June 2025 and July 2025.

### Reasons for ineligibility and missed cases

Reasons for ineligibility were documented for 69 (75.0%) of the 92 ineligible cases (Table 4); the remaining 23 arose during the early implementation phases and during the peak mortality period of June and July 2025, when case volume outstripped the capacity of the monthly summary reporting system. The two operationally significant categories were: 1) in-hospital deaths (27 cases, 6.6%) and 2) deaths notified beyond the 48-hour window (26 cases, 6.4%) which was most prevalent in the early pilot phase before the incentivised informant network was fully established. A further 25 cases (13 in January; 8 in February and 4 in mid-March 2026) were eligible by all criteria but could not proceed due to MITS commodity stockouts. This was the single largest point of disruption in the study, during which all 25 families of deceased individuals were engaged and willing to provide consent. As this was an operational barrier rather than a criterion-based exclusion, these cases were counted as eligible-but-not-enrolled deaths (Table 5). They were formerly reported as “missed eligible” rather than ineligible.

**Table 4.**
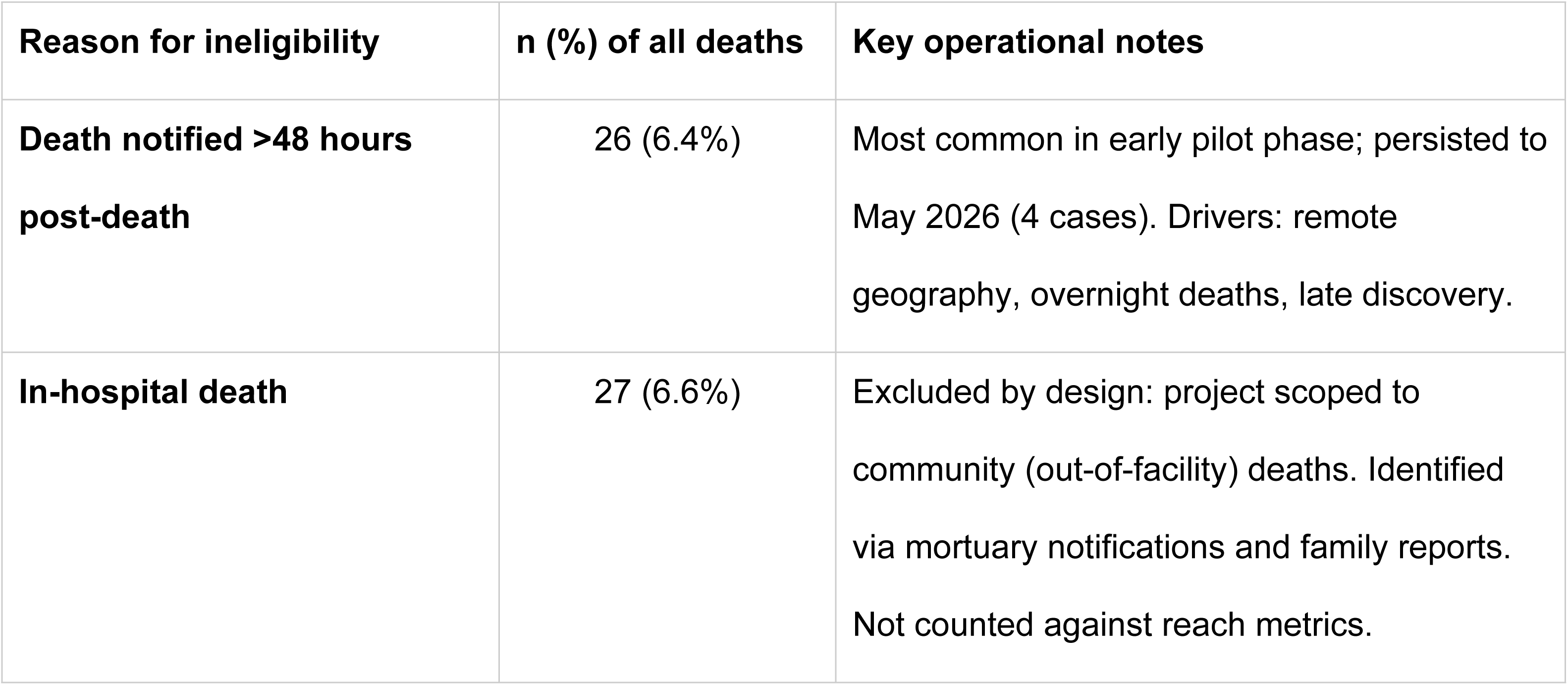
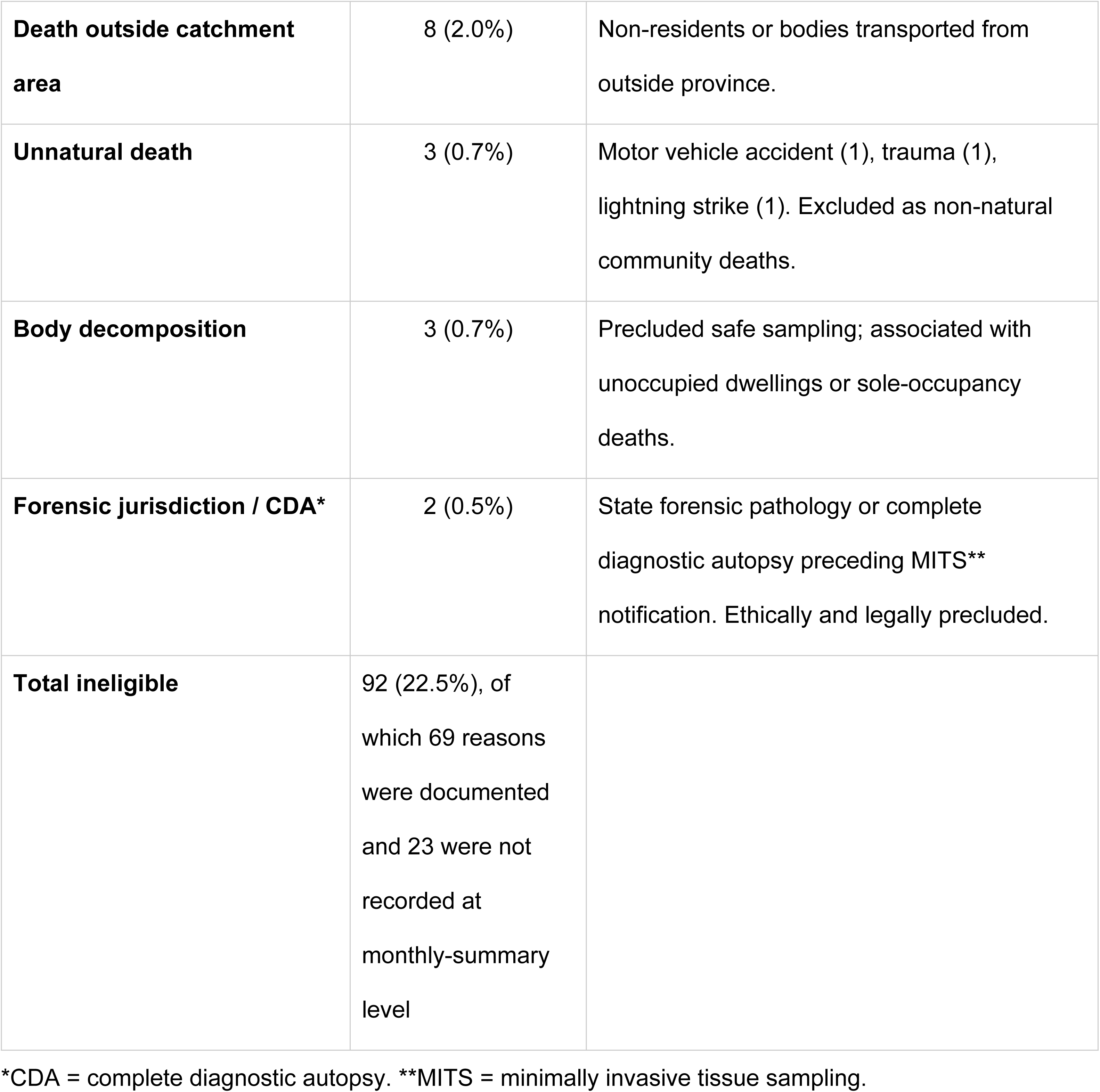
Reasons for MITS ineligibility in the Vukamberi Project, July 2024 – June 2026.

| **Reason for ineligibility** | **n (%) of all deaths** | **Key operational notes** |
|---|---|---|
| **Death notified >48 hours post-death** | 26 (6.4%) | Most common in early pilot phase; persisted to May 2026 (4 cases). Drivers: remote geography, overnight deaths, late discovery. |
| **In-hospital death** | 27 (6.6%) | Excluded by design: project scoped to community (out-of-facility) deaths. Identified via mortuary notifications and family reports. Not counted against reach metrics. |
| **Death outside catchment area** | 8 (2.0%) | Non-residents or bodies transported from outside province. |
| **Unnatural death** | 3 (0.7%) | Motor vehicle accident (1), trauma (1), lightning strike (1). Excluded as non-natural community deaths. |
| **Body decomposition** | 3 (0.7%) | Precluded safe sampling; associated with unoccupied dwellings or sole-occupancy deaths. |
| **Forensic jurisdiction / CDA\*** | 2 (0.5%) | State forensic pathology or complete diagnostic autopsy preceding MITS** notification. Ethically and legally precluded. |
| **Total ineligible** | 92 (22.5%), of which 69 reasons were documented and 23 were not recorded at monthly-summary level |  |

\*CDA = complete diagnostic autopsy. \*\*MITS = minimally invasive tissue sampling.

**Table 5.**
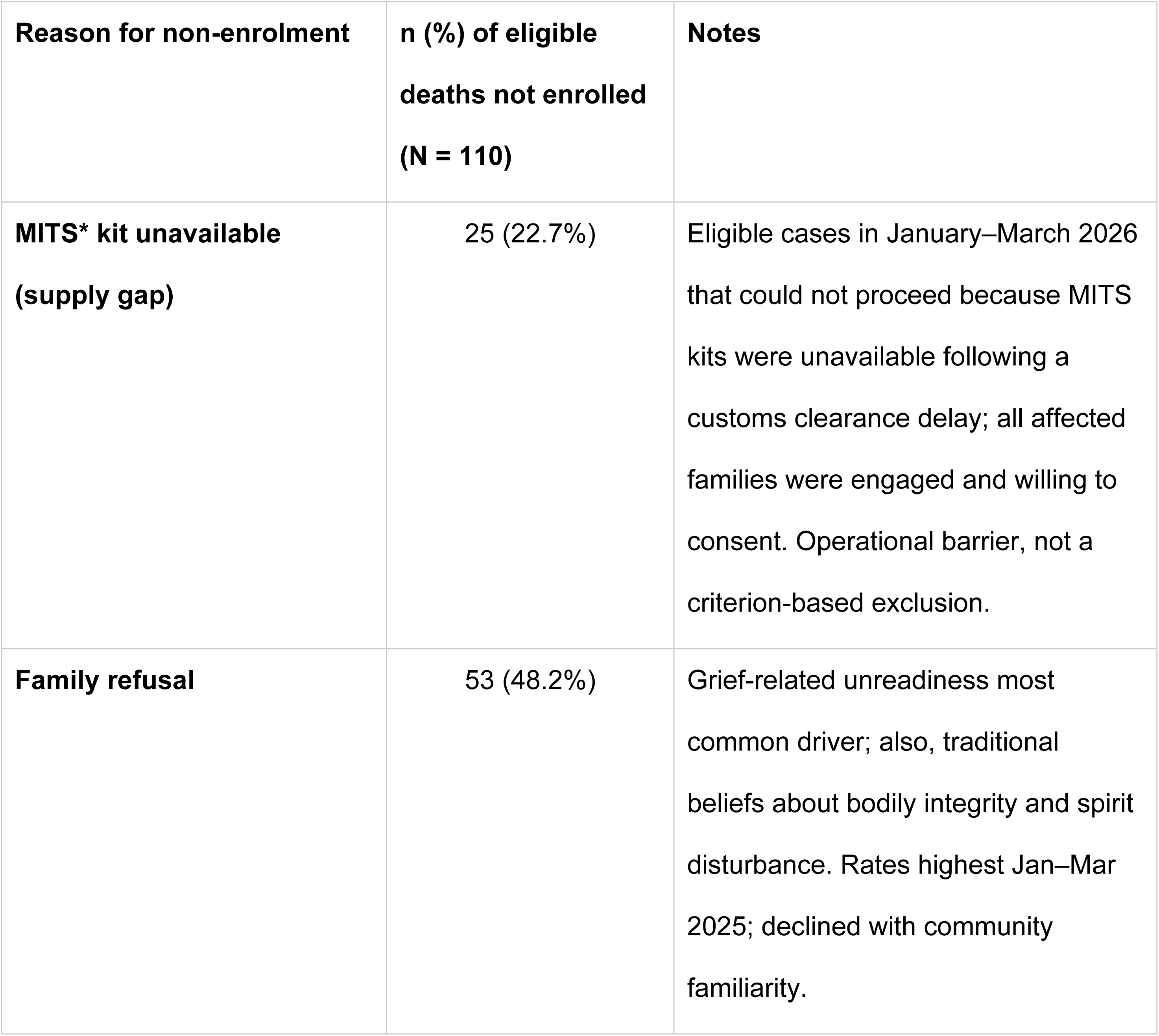
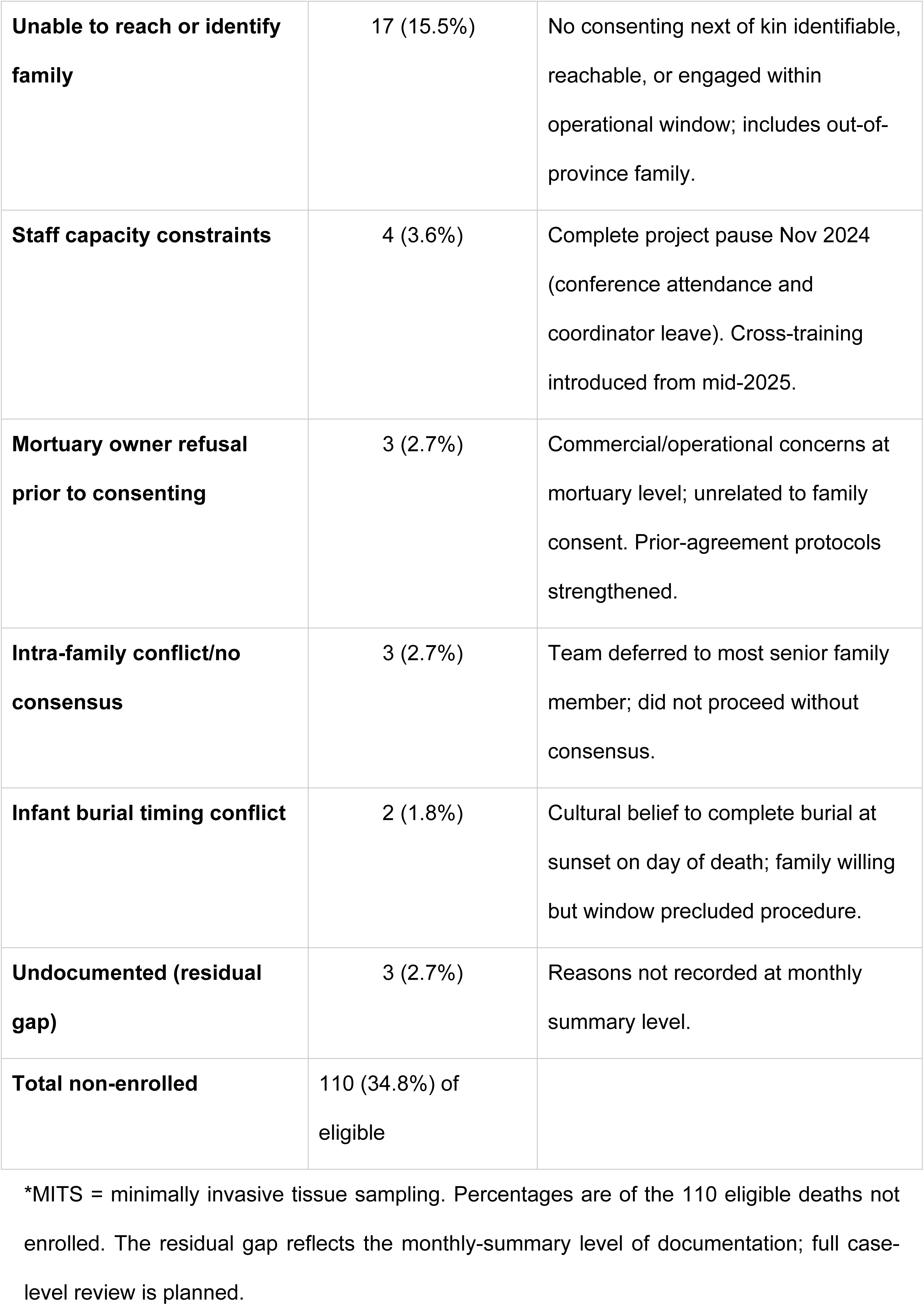
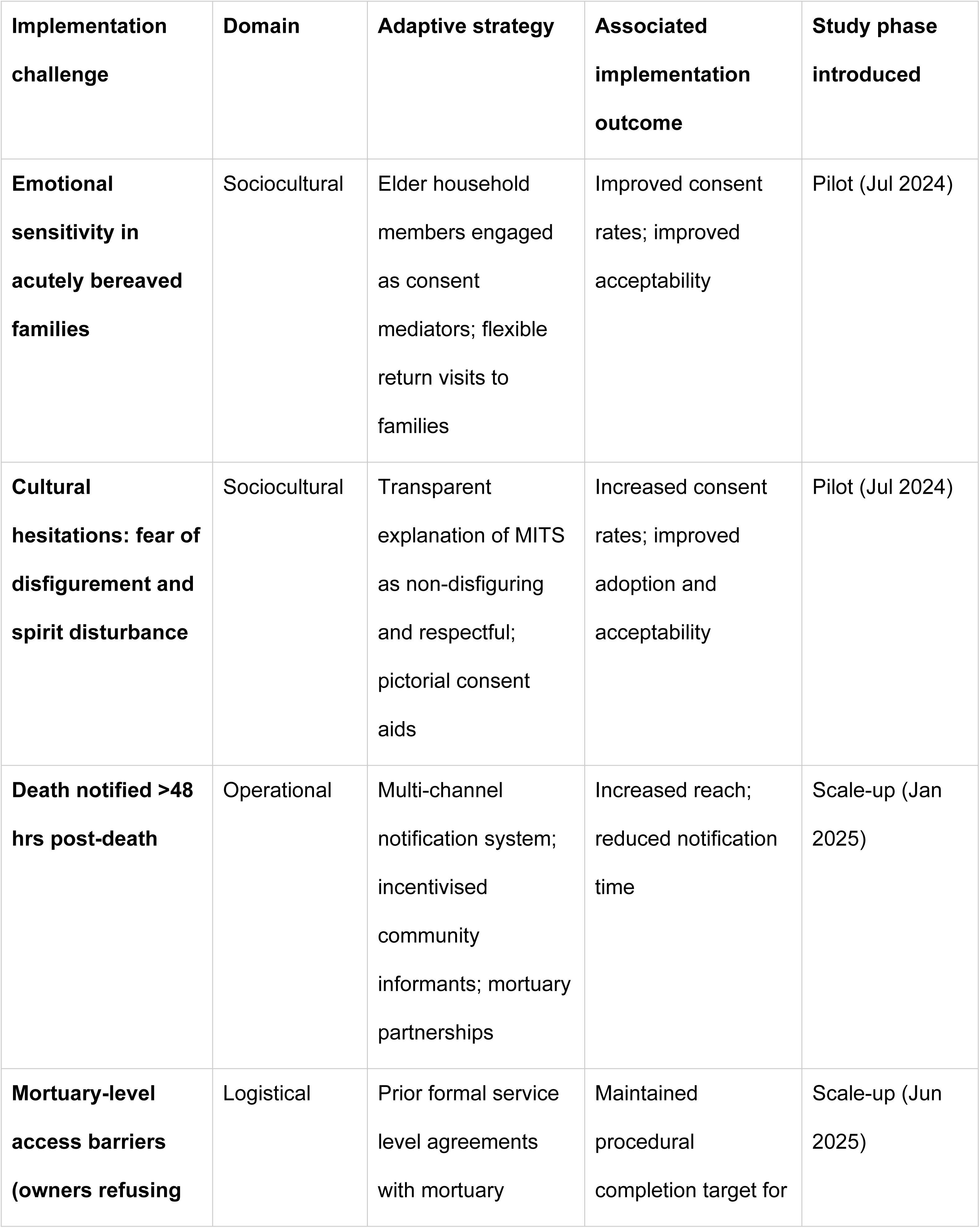
Reasons for non-enrolment among eligible cases, Bushbuckridge sub-district, July 2024 – June 2026.

| Reason for non-enrolment | n (%) of eligible deaths not enrolled (N = 110) | Notes |  |  |
|---|---|---|---|---|
| **MITS\* kit unavailable (supply gap)** | 25 (22.7%) | Eligible cases in January–March 2026 that could not proceed because MITS kits were unavailable following a customs clearance delay; all affected families were engaged and willing to consent. Operational barrier, not a criterion-based exclusion. |  |  |
| **Family refusal** | 53 (48.2%) | Grief-related unreadiness most common driver; also, traditional beliefs about bodily integrity and spirit disturbance. Rates highest Jan–Mar 2025; declined with community familiarity. |  |  |
| **Unable to reach or identify family** | 17 (15.5%) | No consenting next of kin identifiable, reachable, or engaged within operational window; includes out-of-province family. |  |  |
| **Staff capacity constraints** | 4 (3.6%) | Complete project pause Nov 2024 (conference attendance and coordinator leave). Cross-training introduced from mid-2025. |  |  |
| **Mortuary owner refusal prior to consenting** | 3 (2.7%) | Commercial/operational concerns at mortuary level; unrelated to family consent. Prior-agreement protocols strengthened. |  |  |
| **Intra-family conflict/no consensus** | 3 (2.7%) | Team deferred to most senior family member; did not proceed without consensus. |  |  |
| **Infant burial timing conflict** | 2 (1.8%) | Cultural belief to complete burial at sunset on day of death; family willing but window precluded procedure. |  |  |
| **Undocumented (residual gap)** | 3 (2.7%) | Reasons not recorded at monthly summary level. |  |  |
| **Total non-enrolled** | 110 (34.8%) of eligible |  |  |  |
| **Implementation challenge** | **Domain** | **Adaptive strategy** | **Associated implementation outcome** | **Study phase introduced** |
| **Emotional sensitivity in acutely bereaved families** | Sociocultural | Elder household members engaged as consent mediators; flexible return visits to families | Improved consent rates; improved acceptability | Pilot (Jul 2024) |
| **Cultural hesitations: fear of disfigurement and spirit disturbance** | Sociocultural | Transparent explanation of MITS as non-disfiguring and respectful; pictorial consent aids | Increased consent rates; improved adoption and acceptability | Pilot (Jul 2024) |
| **Death notified >48 hrs post-death** | Operational | Multi-channel notification system; incentivised community informants; mortuary partnerships | Increased reach; reduced notification time | Scale-up (Jan 2025) |
| **Mortuary-level access barriers (owners refusing** | Logistical | Prior formal service level agreements with mortuary | Maintained procedural completion target for | Scale-up (Jun 2025) |

\*MITS = minimally invasive tissue sampling. Percentages are of the 110 eligible deaths not enrolled. The residual gap reflects the monthly-summary level of documentation; full case-level review is planned.

### Consent and enrolment

Of the 316 eligible deaths, families of 206 (65.2%) provided informed consent, and a MITS procedure was completed in all 206 consented cases (100%); the quarterly cumulative cascade of notification, eligibility and enrolment is given in Table 3. Field workers recorded the family’s reasons at every refused consent encounter, and these were reviewed at weekly team meetings from the pilot phase onward. These reviews led to the elder-mediated consent, transparent non-disfigurement messaging, pictorial consent aids and flexible return visits that were in place before scale-up began (Table 6). The enrolled series comprised 197 adults (95.6%) and 9 paediatric cases (4.4%, aged 4–16 years); two cases had incomplete age documentation but were confirmed as adults on secondary review. No stillbirths, neonatal deaths, or infant deaths were enrolled during the study period. The protocol was open to all ages from inception; the first paediatric case was enrolled in February 2025. Infant deaths did occur in the catchment: two eligible infant deaths were notified but could not proceed because the families planned burial at sunset on the day of death (Table 5). The absence of infant enrolments therefore points to lower community acceptability of MITS for infant deaths and to same-day burial practice for infants. Elder household members were involved in consent discussions in 277/316 (87.7%) of eligible cases. Elder involvement was associated with improved family receptiveness in field notes where initial hesitancy or intra-family disagreement had been documented. The complete enrolment cascade is presented in Fig 4.

**Fig 4.**
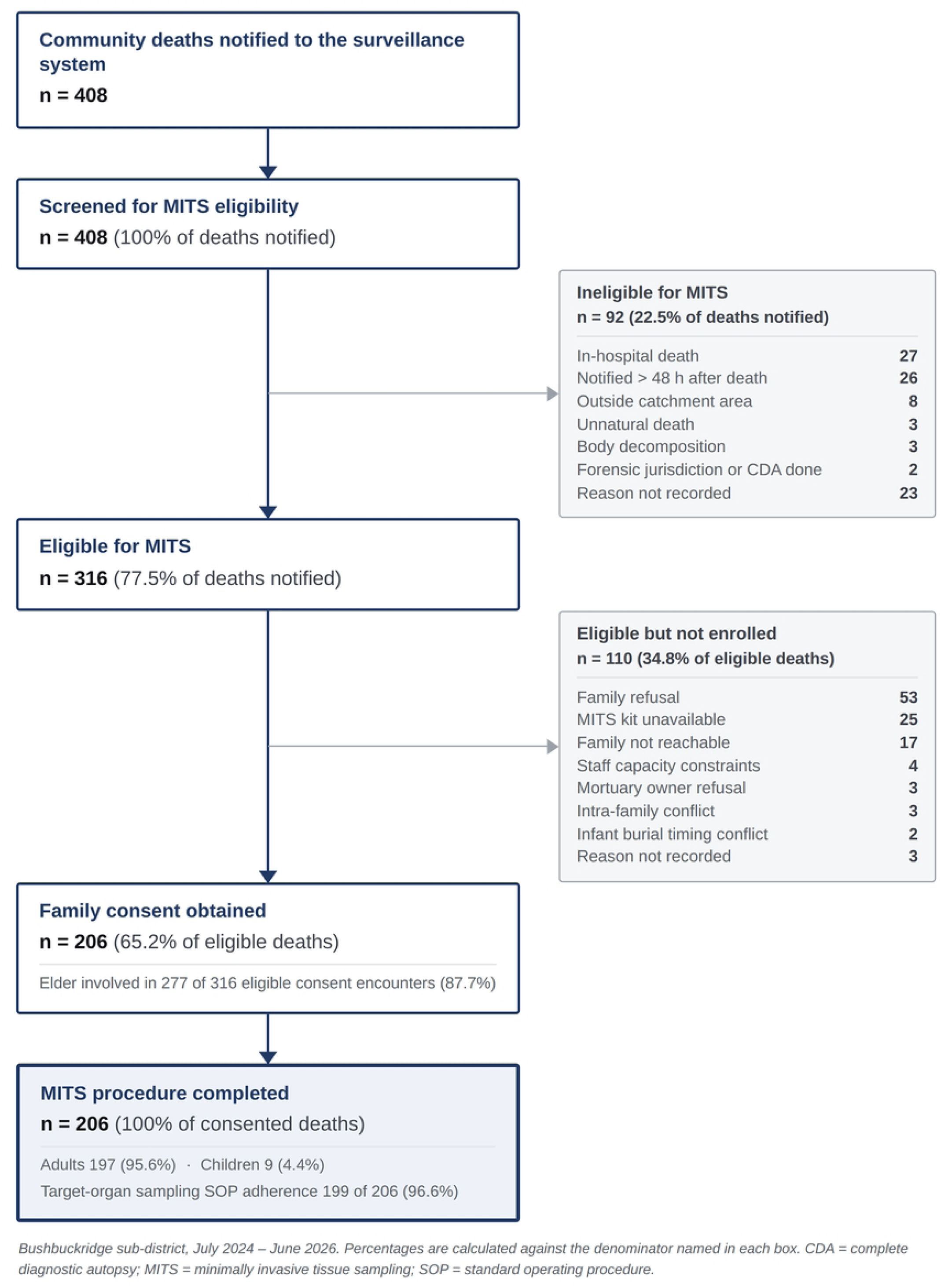
Vukamberi enrolment cascade, July 2024 – June 2026.

**Table 6.** Implementation challenges and adaptive strategies by domain, The Vukamberi Project, July 2024 – June 2026.

| Implementation challenge | Domain | Adaptive strategy | Associated implementation outcome | Study phase introduced |
|---|---|---|---|---|
| **Emotional sensitivity in acutely bereaved families** | Sociocultural | Elder household members engaged as consent mediators; flexible return visits to families | Improved consent rates; improved acceptability | Pilot (Jul 2024) |
| **Cultural hesitations: fear of disfigurement and spirit disturbance** | Sociocultural | Transparent explanation of MITS as non-disfiguring and respectful; pictorial consent aids | Increased consent rates; improved adoption and acceptability | Pilot (Jul 2024) |
| **Death notified >48 hrs post-death** | Operational | Multi-channel notification system; incentivised community informants; mortuary partnerships | Increased reach; reduced notification time | Scale-up (Jan 2025) |
| **Mortuary-level access barriers (owners refusing entry; family approach and burial timing conflicts)** | Logistical | Prior formal service level agreements with mortuary partners; team schedule adjusted to avoid peak mortuary operations | Maintained procedural completion target for consented cases and timeliness | Scale-up (Jun 2025) |
| **Local chief restricting community entry in 14 villages** | Sociopolitical | Families approached at mortuaries for consent outside the chief's jurisdiction | Increased reach; improved adoption | Scale-up (Aug 2025) |
| **Laboratory turnaround delays** | Project-level | In-house culture bench established (Jan 2026); prioritised laboratory workflow | Improved sample adequacy; reduced turnaround time and costs | Full surveillance (Jan 2026) |
| **MITS kit supply gap (Jan–Mar 2026)** | Operational / supply chain | Emergency kit procurement; enhanced stock monitoring and buffer inventory from Apr 2026 | 21 cases missed (Jan–Feb 2026); 4 cases missed (Mar 2026); supply restored | Full surveillance (Jan 2026) |

MITS = minimally invasive tissue sampling; SOP = standard operating procedure. Challenges are mapped to the implementation phase and domain in which they were first identified. Adaptive strategies and associated outcomes reflect documented field team responses.

### Reasons for non-enrolment among eligible cases

Family refusal (53 cases, 48.2% of non-enrolled eligible cases) was the single largest reason for non-enrolment, documented in 18 of the 24 months of the study. Reasons for refusal were recorded at monthly-summary level without the age or sex of the decedent, so variation in refusal by age or sex could not be assessed. Refusals were heterogeneous (Table 5): the most common driver was grief-related unreadiness, where families in the acute aftermath of bereavement were not yet able to engage with the consent process. Traditional beliefs about bodily integrity and disturbance of the spirit of the deceased were documented in a subset of cases. In several instances, families expressed fatalistic acceptance of the death particularly for elderly decedents as in one field-documented case where the family stated the deceased “was old, they had accepted the outcome and did not want her to be part of research.” In two cases, families explicitly declined further contact. Monthly refusal rates were highest during the early scale-up period (January–March 2025: 5 per month) and, apart from a return to five in June 2025 (the peak enrolment month), remained at four or fewer per month thereafter as community familiarity grew. The monthly refusal trend is overlaid on Fig 5. The concentration of refusals in the months immediately following the extension of enrolment into newly engaged villages suggests that community sensitisation should be completed before enrolment begins in a new area.

**Fig 5.**
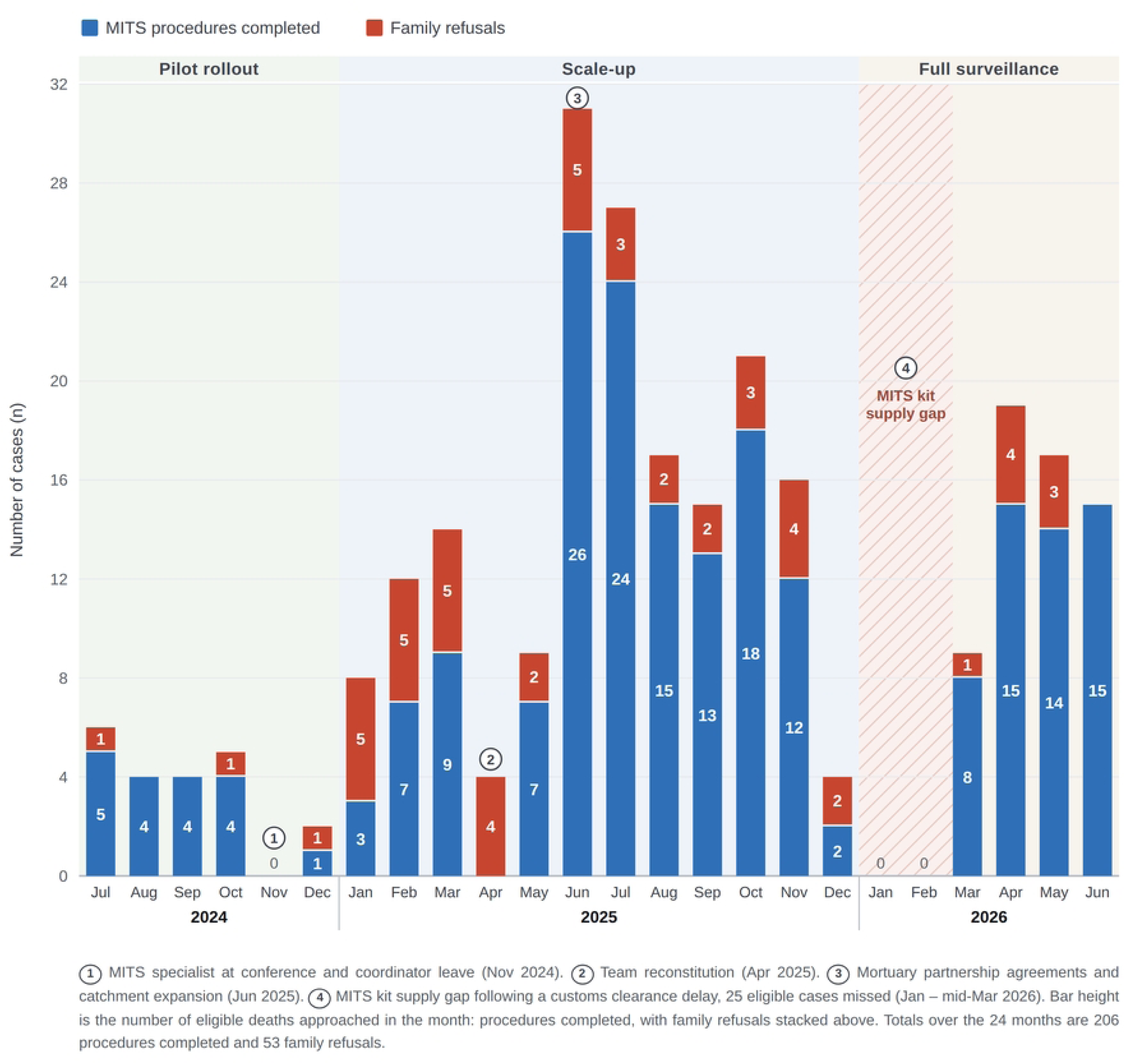
Monthly MITS enrolment and family refusals with implementation phases and adaptations, Bushbuckridge sub-district, July 2024 – June 2026 (N = 206).

### Enrolment trajectory and programme ramp-up

Monthly MITS enrolment followed the three implementation phases of the enrolment period (pilot, scale-up and full surveillance), summarised in Table 1. During the pilot phase (July– December 2024) enrolments ranged from 0–5 per month; the sole zero-enrolment month (November 2024) coincided with the MITS specialist’s conference attendance and coordinator leave. Within the scale-up phase, enrolment accelerated progressively to a peak of 26 procedures in June 2025 following a brief operational pause in April 2025 for team reconstruction. The June peak may have been amplified by higher winter mortality which coincided with full operationalisation of the expanded notification network and mortuary partnerships. The adaptive strategies underpinning these gains, and the study phase in which each was introduced, are summarised in Table 6. Enrolment stabilised at 12–18 procedures per month (August–November 2025) before the kit supply disruption arising from a customs clearance delay paused enrolments in January–February 2026. Recovery from stockouts was rapid: 8 cases in March, 15 cases in April, 14 cases in May and 15 cases in June 2026. Median monthly enrolment across the full period was 7.5 procedures (range 0–26). The monthly enrolment trajectory with study phase demarcations is presented in Fig 5.

### Procedural completion and sample collection

All enrolled participants had tissue and, where obtainable, fluid samples successfully collected in accordance with the MITS standard operating procedures. Specimen collection was near-complete for the principal diagnostic compartments. Among the 206 enrolled cases, formalin-fixed tissue was obtained from the brain in 203 (98.5%), right lung in 205 (99.5%), left lung 204 (99.0%) and the liver in 204 (99.0%); the four core solid organs were jointly sampled in 201 cases (97.6%). Kidney tissue was obtained in a minority of cases (left 29 [14.1%], right 24 [11.7%]); kidney biopsy was attempted for all cases but was frequently constrained by available field time relative to the core diagnostic organs. For microbiology, EDTA whole blood was collected in 203 cases (98.5%), blood culture bottles (aerobic and anaerobic) in 201 cases (97.6%), nasopharyngeal swabs in 205 (99.5%; collected but not yet tested, owing to cost constraints), and cerebrospinal fluid (CSF) in 180 (87.4%), with paired solid-organ microbiology specimens obtained in 200 cases (97.1%). Rectal swabs were not collected. The study team decided against them when developing the protocol because the budget could not cover the multiplex molecular testing needed to distinguish enteric pathogens that contributed to death from normal gut flora. Sample adequacy for the core specimen set was therefore high.

### Sample processing and analysis

The collected specimens are distinct from completed analyses with laboratory processing, verbal autopsies, medical records linkage and determination of cause of death (DeCoDe) processes continuing. As of June 2026, microbiological processing on the in-house culture bench had been finalised for 108 of 206 cases (52.4%). The outstanding cases reflect laboratory throughput at the reporting cut-off, and processing was continuing at the close of the study period. Procedural completion tracked cumulative eligibility across the study quarters (Table 3). Histology samples were submitted for 198 cases, of which 100 (50.5%) have been completed. VA and TeleVA interviews were completed for 143 of 206 enrolled cases (69.4%). Clinical records from the Clinic-link HDSS project include ante-mortem laboratory results, diagnosis, medication, clinic and hospital visits. Records were retrieved for 50 of the 92 decedents residing within the HDSS at the time of death (54.3%; 24.3% of all 206 enrolled cases); no Clinic-link record exists for the remaining 114 decedents, who were resident beyond the HDSS (106) or were former residents of the study area (8). DeCoDe multi-disciplinary CoD panel determinations have been finalised for 100/206 (48.5%). Full laboratory analysis and CoD findings will be reported in a dedicated epidemiological manuscript.

The laboratory analysis status by assay is summarised in Fig 6. The five workstreams are not equally advanced, and the reasons differ: histology depends on an external referral laboratory and its turnaround, verbal autopsy depends on a return visit to a bereaved household and is deliberately deferred to respect the culturally accepted mourning period, and the DeCoDe panel cannot convene on a case until all results from a given case are available. Clinic-link retrieval was limited by design, because, ante-mortem records exist only for decedents resident within the HDSS catchment; for the remaining decedents no record can be linked, and these cases are shown separately in Fig 6 rather than counted as outstanding (50 retrieved, 42 pending and 114 not linkable).

**Fig 6.**
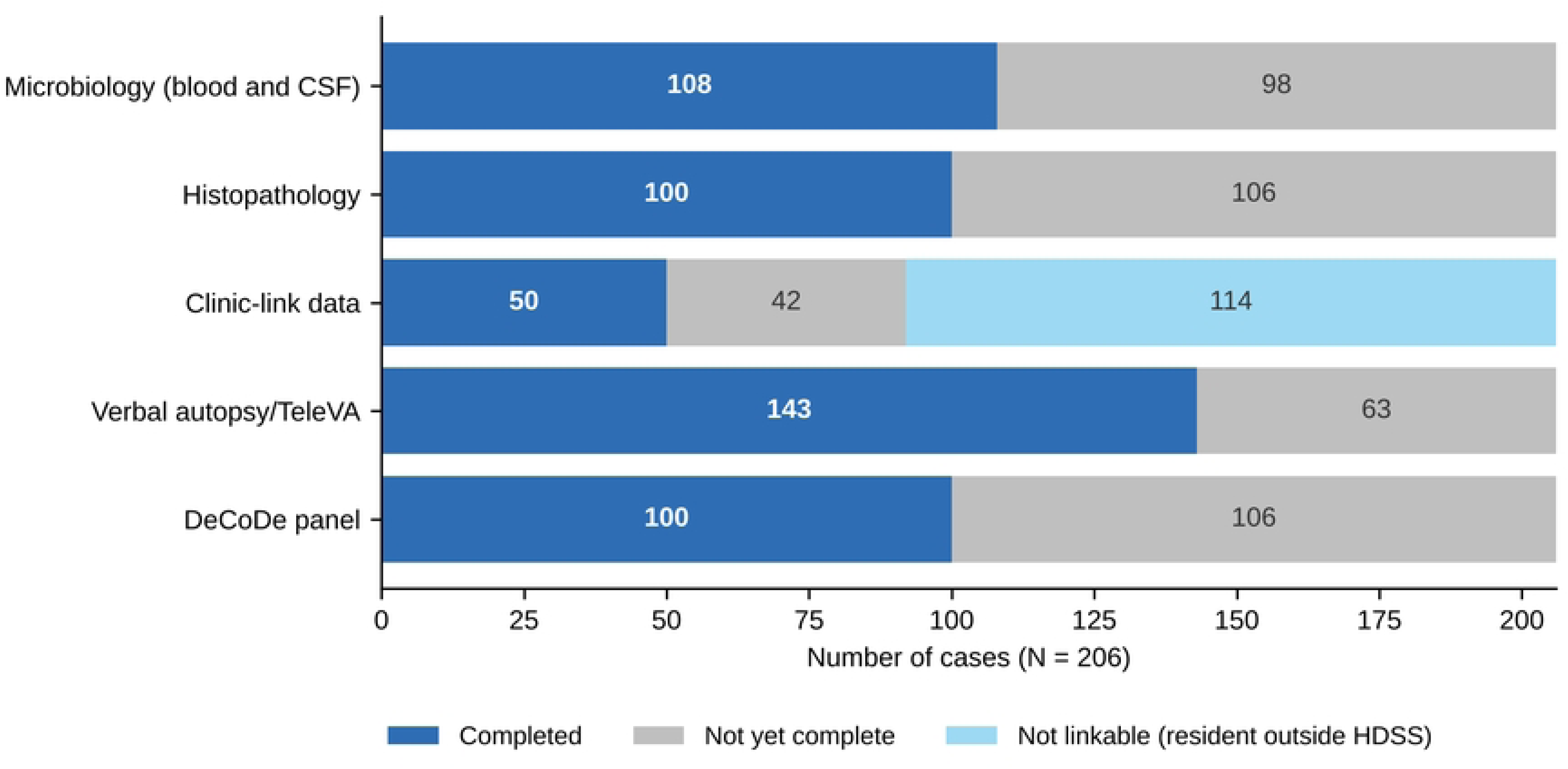
Laboratory analysis status by assay type as of June 2026.

Point-of-care (POC) testing performed at the time of MITS sampling, was completed in 205 of 206 enrolled cases (99.5%). Detailed serological findings will be reported in a dedicated epidemiological manuscript.

### Implementation challenges and the adaptive strategies introduced

Drawing the domains together, implementation across the four phases was shaped by a recurring set of challenges: notification arriving more than 48 hours after death, emotional sensitivity in acutely bereaved families, cultural hesitancy about disfigurement and disturbance of the spirit of the deceased, mortuary-level access barriers, a traditional leader’s restriction on entry to 14 villages, laboratory turnaround delays, and the MITS kit supply gap of January to mid-March 2026. Each was met with an adaptation introduced at an identifiable point in the study and traceable to a change in the implementation outcomes reported above: the incentivised multi-channel notification network restored reach and reduced out-of-window notification; elder mediation and transparent non-disfigurement messaging raised consent; formal mortuary agreements and mortuary-based consenting preserved reach and procedural completion where household access failed; and the in-house culture bench shortened laboratory turnaround. Taken together these adaptations describe an implementation model in which community-based MITS is sustained by the notification, consent and laboratory systems built around the sampling procedure (Table 6).

## Discussion

### Principal findings

This study presents, to our knowledge, one of the first systematic process evaluations of establishing community-based all-ages MITS surveillance across all ages. Over 24 months of enrolment, all 408 notified community deaths were screened, a MITS procedure was completed in all 206 consented cases with target-organ sampling adherence of 96.6%, and 65.2% of eligible families consented, indicating that community-based MITS is operationally feasible within an established rural surveillance platform. Enrolment increased from fewer than five procedures per month in the pilot phase to 12–26 per month from June to November 2025 and recovered to 14–15 per month from April 2026 after the kit supply gap, consistent with iterative adaptation to context-specific barriers within the HDSS infrastructure [24,26,27]. The evaluation also extends the MITS evidence base: CHAMPS and most published MITS programmes have focused on stillbirths and child deaths, whereas Vukamberi applied the procedure to community deaths at all ages, including adults. The diagnostic validity of MITS against complete diagnostic autopsy was established in hospital deaths in Mozambique, with cause-of-death concordance of 75.9% in adults, 89% in children, 83% in stillbirths, 68% in neonates and 68% in maternal deaths [30–33]; whether that performance holds for community deaths sampled a median of 37 hours, and up to 72 hours, after death will be addressed by the Vukamberi cause-of-death analysis.

### Death notification and timeliness

Screening of all notified deaths was maintained throughout the study, although this measures coverage of notified deaths rather than of all deaths in the catchment (see Limitations). CHAMPS and comparable programmes typically target notification within 24–72 hours for paediatric deaths in both communities and health facilities [16]. Our 48-hour window for all-age community deaths was chosen because high ambient temperatures in the study area accelerate body decomposition and bacterial overgrowth, and because of the culturally grounded practice among elders of waiting two to six hours after apparent death before calling the mortuary, which is linked to beliefs of possible ‘miraculous recovery’ [24,34]. Only 26 deaths (6.4%) were ineligible because of delayed notification, a pattern consistent with community-based MITS programmes in other rural settings, including India and Pakistan, where cultural practices around death disclosure, geographic isolation and limited informant awareness contribute to delayed reporting [19,20]. The multi-channel notification network, particularly incentivised community and mortuary informants, was the main strategy that sustained screening and reduced out-of-window notifications across study phases.

### Consent, refusal and elder mediation

The 65.2% consent rate among eligible cases is lower than the 82% consent reported for paediatric deaths in a Mozambican hospital setting [17] but was achieved for community deaths of all ages within a 48-hour window and is consistent with the high hypothetical acceptability of MITS reported in this community during the Vukamberi formative study [24]. Qualitative studies on MITS implementation from Kenya, Mozambique and rural South Africa identified fears of organ harvesting, spirit disturbance and burial delay as the main barriers to consent; all three were documented in Vukamberi field notes and addressed through transparent communication and elder mediation [18,24,34]. The Kenyan acceptability study similarly found that community health workers and elder community figures were key facilitators, supporting the transferability of elder-mediated consent across sub-Saharan African settings [18]. Recent MITS implementation in Zambezia Province, central Mozambique, also found that advocacy involving provincial government, community leaders and traditional healers was critical for sustained acceptability [35].

The dominant driver of non-enrolment was family refusal (48.2% of non-enrolled eligible cases), most often reflecting grief-related unreadiness. This pattern aligns with evidence from consent research in acute bereavement contexts, which identifies emotional state at the time of approach as a primary determinant of participation, independent of attitudes toward the study itself [36]. Most refusals reflected a mismatch between the 48-hour operational window and a family’s capacity to engage in the consent process, and few families expressed principled opposition to MITS. This has implications for programme design: advance community sensitisation that allows households to indicate openness to participation before bereavement, and flexible return-visit protocols, may increase enrolment without compromising consent. The decline in refusals as community familiarity grew across phases supports this interpretation, although refusal reasons were recorded at summary level and could not be linked to decedent characteristics.

A distinctive feature of Vukamberi implementation was the strategic use of elder household members as consent mediators. Our earlier qualitative study in this community documented how elder authority in Xitsonga- and Sepulana (Northern Sotho)-speaking communities in rural Mpumalanga is deeply embedded in death ritual governance; elders hold decision-making power over whether the deceased’s body may be examined, and their endorsement is both socially required and practically decisive for family consent decisions [34]. This authority is not static: the same study found that younger household members increasingly defer to elders on decisions made around the time of death, such as whether the body may be examined, even in households where elders have less say in everyday matters. Elder involvement was documented in 277/316 (87.7%) of eligible cases and was consistently associated with improved family receptiveness in field notes where initial hesitancy was documented. This finding is consistent with evidence from the CHAMPS PICK-CHAMP participatory community- entry workshops, in which participants across five sites indicated that support for MITS was contingent on the procedure not interfering with burial practices and rituals, and in which engagement with community leaders was central to establishing acceptability [37]. The sociopolitical barrier of local chief’s access restriction in 14 villages was managed through mortuary-based consenting, an adaptation that preserved reach by drawing on a structural feature of rural South African mortuary practice, in which families are reliably present in the peri-death period. A comparable emphasis on rapid identification of deaths and on prior institutional relationship-building was reported from Zambezia Province, central Mozambique, where these were identified as preconditions for timely sampling [35].

### Procedural fidelity and supply-chain vulnerability

Target-organ sampling adherence of 96.6% (199/206 consented cases) indicates high procedural fidelity, supported by established mortuary partnerships and consistent team training. The 48-hour eligibility window was set on operational grounds, reflecting ambient temperature, the absence of refrigeration in most households and local burial practice, rather than on a published post-mortem interval threshold. The most significant operational disruption was the MITS kit supply gap from January to mid-March 2026, which resulted in 25 missed eligible cases at a time when families were willing to consent. Cost-evaluation work in LMICs has identified MITS kits and consumables, alongside labour and testing, as principal cost drivers [38]; our experience shows that they are also a key operational vulnerability and underscores the need for buffer stock and procurement redundancy as projects move from externally funded pilots to integrated surveillance systems. Because the scientific value of surveillance depends on population-representative sampling, such gaps introduce non-random missingness: a seasonal block of community deaths is absent from the series for reasons unrelated to the characteristics of those deaths.

### Point-of-care testing

Point-of-care (POC) testing was performed at the time of MITS sampling in 205 of 206 cases (99.5%). Integrating POC testing for HIV, hepatitis B surface antigen, hepatitis C antibody and syphilis (RPR/TPHA) into the MITS procedure adds minimal procedural time and provides immediate serological context directly relevant to interpreting cause-of-death pathways, which may be useful for sites without rapid access to laboratory serology.

### Implications for implementation and policy

Viewed through an implementation science lens, the Vukamberi trajectory illustrates core principles of effective complex intervention implementation: staged rollout with deliberate preparatory investment prior to procedures; iterative adaptation in response to identified barriers; and system-level capacity building, including in-house culture bench establishment and mortuary partnership formalisation that improved project performance over time [26]. The preparatory phase (May–December 2023), focused on community engagement, stakeholder mapping, and formative acceptability work, was important for building the social licence necessary to conduct post-mortem procedures in bereaved households in a context where mistrust of biomedical research remains a real concern. Our Phase 1 qualitative study reported that MITS was broadly acceptable in this community, with community members expressing willingness to participate provided the procedure was non-disfiguring, respectfully conducted, and that results would be shared back with families and the community; concerns about spirit disturbance and organ harvesting were raised and were specifically addressed through the pictorial consent aids and community information sessions developed as part of that preparatory phase [24]. This mirrors the PICK-CHAMP community-entry model, in which structured pre-implementation engagement was used to surface alignment and tension between surveillance activities and community priorities before mortality surveillance began [37]. The Africa CDC Continental Framework for Mortality Surveillance explicitly calls for integration of community mortality surveillance into national health information systems; the Vukamberi experience offers an operational example of how this could be done at district level [39].

### Strengths and limitations

This study has several strengths. First, the evaluation integrated MRC process evaluation guidance with Proctor’s Implementation Outcomes Framework, allowing implementation outcomes to be linked to specific mechanisms and contextual factors; to our knowledge, it is among the first systematic process evaluations of community-based MITS and the first to cover deaths at all ages within an HDSS. Second, implementation outcomes were derived from records generated prospectively as the intervention ran (notification logs, consent documentation, field and case reports and laboratory quality records) rather than reconstructed retrospectively, so barriers and the adaptations made in response are dated and can be read against the performance indicators of the period in which they occurred. Third, reporting follows the Standards for Reporting Implementation Studies (StaRI) statement (S1 Checklist) [40].

This evaluation also has limitations. First, implementation outcomes were abstracted from monthly summary reports rather than case-level records, so reasons were not documented for 23 ineligible and 3 non-enrolled cases, and refusals could not be stratified by decedent age, sex or duration of terminal illness; these should be captured routinely in future surveillance. Second, study notifications were not compared with deaths recorded independently by the HDSS, so the completeness of death ascertainment, overall and by age group (including infant deaths), could not be quantified; screening coverage refers to notified deaths only, and ascertainment was incomplete in February 2026, when only two mortuaries reported. Third, the traditional leader’s restriction of access to 14 villages and the kit supply gap of early 2026 may have introduced selection bias into the enrolled series, which may affect the representativeness of future cause-of-death estimates. Fourth, the enrolled series was predominantly adult and included no stillbirths, neonates or infants, so feasibility for these groups could not be assessed. Fifth, cause-of-death determination was incomplete at the reporting cut-off, so the diagnostic yield of community-based MITS is not reported here. Sixth, the evaluation was conducted by the implementing team, which may have influenced the interpretation of field notes; the use of prospectively recorded operational data reduces, but does not remove, this risk. Finally, the single-site design limits generalisability, although the Agincourt setting shares structural features with many surveillance sites in sub-Saharan Africa, including rural geography, high HIV burden, incomplete civil registration and strong community leadership.

## Conclusion

The Vukamberi project demonstrates that community-based MITS surveillance is feasible, operationally achievable, and generates actionable scientific data within an established HDSS in rural South Africa. Elder-mediated consent, mortuary partnerships, multi-channel incentivised death notification and in-house laboratory capacity may be transferable to sites facing comparable barriers in the African region, and reliable procurement of consumables is a precondition for continuous surveillance. Africa CDC’s Continental Framework for Mortality Surveillance calls for community mortality surveillance to be integrated into national health information systems [39]; the Vukamberi experience illustrates what such integration requires in practice. Cause-of-death findings from the complete cohort will provide a pathology-informed community mortality dataset for rural South Africa, offering a resource for validating verbal autopsy, informing public health priorities and tracking the epidemiological transition in a setting of growing non-communicable disease burden.

## Data Availability

The minimal de-identified dataset underlying the implementation outcomes reported here is provided in S1 Data. Individual-level decedent records cannot be shared publicly as they contain potentially identifying information about decedents and bereaved households within a well-defined rural population, and due to participant consent and the governing ethics approvals do not permit open release. Requests for access to these restricted data may be sent to the SAMRC/Wits Agincourt Unit Data Access Committee, which reviews requests independently of the authors.

## Acknowledgments

The authors thank the communities of the Bushbuckridge sub-district for their trust and participation; the Vukamberi field team for their dedication across all implementation phases; the mortuary partners and community informants whose networks made real-time death notification possible; the Public Engagement Office (PEO) and the SAMRC/Wits Rural Public Health and Health Transitions Research Unit (Agincourt) and its Community Advisory Board for guidance throughout. We acknowledge the Nyanja Health Research Institute for teaching in advanced scientific writing, and MITS Surveillance Alliance at RTI for technical guidance and training opportunities.

## Supporting information

**S1 Appendix. Agincourt Health and Demographic Surveillance System: description of the study setting.**

